# The ANTsX ecosystem for quantitative biological and medical imaging

**DOI:** 10.1101/2020.10.19.20215392

**Authors:** Nicholas J. Tustison, Philip A. Cook, Andrew J. Holbrook, Hans J. Johnson, John Muschelli, Gabriel A. Devenyi, Jeffrey T. Duda, Sandhitsu R. Das, Nicholas C. Cullen, Daniel L. Gillen, Michael A. Yassa, James R. Stone, James C. Gee, Brian B. Avants, for the Alzheimer’s Disease Neuroimaging Initiative

**Affiliations:** Department of Radiology and Medical Imaging, University of Virginia, Charlottesville, VA; Department of Radiology, University of Pennsylvania, Philadelphia, PA; Department of Biostatistics, University of California, Los Angeles, CA; Department of Electrical and Computer Engineering, University of Iowa, Philadelphia, PA; School of Public Health, Johns Hopkins University, Baltimore, MD; Douglas Mental Health University Institute, Department of Psychiatry, McGill University, Montreal, QC; Lund University, Scania, SE; Department of Statistics, University of California, Irvine, CA; Department of Neurobiology and Behavior, University of California, Irvine, CA

## Abstract

The Advanced Normalizations Tools ecosystem, known as ANTsX, consists of multiple open-source software libraries which house top-performing algorithms used worldwide by scientific and research communities for processing and analyzing biological and medical imaging data. The base software library, ANTs, is built upon, and contributes to, the NIH-sponsored Insight Toolkit. Founded in 2008 with the highly regarded Symmetric Normalization image registration framework, the ANTs library has since grown to include additional functionality. Recent enhancements include statistical, visualization, and deep learning capabilities through interfacing with both the R statistical project (ANTsR) and Python (ANTsPy). Additionally, the corresponding deep learning extensions ANTsRNet and ANTsPyNet (built on the popular TensorFlow/Keras libraries) contain several popular network architectures and trained models for specific applications. One such comprehensive application is a deep learning analog for generating cortical thickness data from structural T1-weighted brain MRI, both cross-sectionally and longitudinally. These pipelines significantly improve computational efficiency and provide comparable-to-superior accuracy over multiple criteria relative to the existing ANTs workflows and simultaneously illustrate the importance of the comprehensive ANTsX approach as a framework for medical image analysis.

## The ANTsX ecosystem: A brief overview

### Image registration origins

The Advanced Normalization Tools (ANTs) is a state-of-the-art, open-source software toolkit for image registration, segmentation, and other functionality for comprehensive biological and medical image analysis. Historically, ANTs is rooted in advanced image registration techniques which have been at the forefront of the field due to seminal contributions that date back to the original elastic matching method of Bajcsy and co-investigators.^54,55,59^ Various independent platforms have been used to evaluate ANTs tools since their early development. In a landmark paper,^65^ the authors reported an extensive evaluation using multiple neuroimaging datasets analyzed by fourteen different registration tools, including the Symmetric Normalization (SyN) algorithm,^60^ and found that “ART, SyN, IRTK, and SPM’s DARTEL Toolbox gave the best results according to overlap and distance measures, with ART and SyN delivering the most consistently high accuracy across subjects and label sets.” Participation in other independent competitions^62,69^ provided additional evidence of the utility of ANTs registration and other tools.^13,14,42^ Despite the extremely significant potential of deep learning for image registration algorithmic development,^41^ ANTs registration tools continue to find application in the various biomedical imaging research communities.

### Current developments

Since its inception, though, ANTs has expanded significantly beyond its image registration origins. Other core contributions include template building,^64^ segmentation,^68^ image pre-processing (e.g., bias correction^52^ and denoising),^56^ joint label fusion,^53,63^ and brain cortical thickness estimation^57,66^ (cf Table 1). Additionally, ANTs has been integrated into multiple, publicly available workflows such as fMRIprep^50^ and the Spinal Cord Toolbox.^49^ Frequently used ANTs pipelines, such as cortical thickness estimation,^66^ have been integrated into Docker containers and packaged as Brain Imaging Data Structure (BIDS)^48^ and FlyWheel applications (i.e., “gears’ ’). It has also been independently ported for various platforms including Neurodebian^47^ (Debian OS), Neuroconductor^46^ (the R statistical project), and Nipype^45^ (Python). Additionally, other widely used software, such as FreeSurfer,^61^ have incorporated well-performing and complementary ANTs components^52,56^ into their own libraries. According to GitHub, recent unique “clones” have averaged 34 per day with the total number of clones being approximately twice that many. 50 unique contributors to the ANTs library have made a total of over 4500 commits. Additional insights into usage can be viewed at the ANTs GitHub website.

**Table 1:** The significance of core ANTs tools in terms of their number of citations (from October 17, 2020).

| Functionality | Citations |
| --- | --- |
| SyN registration<sup>5</sup> | 2616 |
| bias field correction<sup>16</sup> | 2188 |
| ANTs registration evaluation<sup>6</sup> | 2013 |
| joint label fusion<sup>18</sup> | 669 |
| template generation<sup>14</sup> | 423 |
| cortical thickness: implementation<sup>20</sup> | 321 |
| MAP-MRF segmentation<sup>15</sup> | 319 |
| ITK integration<sup>12</sup> | 250 |
| cortical thickness: theory<sup>19</sup> | 180 |

Over the course of its development, ANTs has been extended to complementary frameworks resulting in the Python- and R-based ANTsPy and ANTsR toolkits, respectively. These ANTs-based packages interface with extremely popular, high-level, open-source programming platforms which have significantly increased the user base of ANTs. The rapidly rising popularity of deep learning motivated further recent enhancement of ANTs and its extensions. Despite the existence of an abundance of online innovation and code for deep learning algorithms, much of it is disorganized and lacks a uniformity in structure and external data interfaces which would facilitate greater uptake. With this in mind, ANTsR spawned the deep learning ANTsRNet package^32^ which is a growing Keras/TensorFlow-based library of popular deep learning architectures and applications specifically geared towards medical imaging. Analogously, ANTsPyNet is an additional ANTsX complement to ANTsPy. Both, which we collectively refer to as “ANTsXNet”, are co-developed so as to ensure cross-compatibility such that training performed in one library is readily accessible by the other library. In addition to a variety of popular network architectures (which are implemented in both 2-D and 3-D), ANTsXNet contains a host of functionality for medical image analysis that have been developed in-house and collected from other open-source projects. For example, an extremely popular ANTsXNet application is a multi-modal brain extraction tool that uses different variants of the popular U-net^44^ architecture for segmenting the brain in multiple modalities. These modalities include conventional T1-weighted structural MRI as well as T2-weighted MRI, FLAIR, fractional anisotropy, and BOLD data. Demographic specialization also includes infant T1-weighted and/or T2-weighted MRI. Additionally, we have included other models and weights into our libraries such as a recent BrainAGE estimation model,^23^ based on > 14, 000 individuals; HippMapp3r,^43^ a hippocampal segmentation tool; the winning entry of the MICCAI 2017 white matter hyperintensity segmentation competition;^40^ MRI super resolution using deep back-projection networks;^22^ and NoBrainer, a T1-weighted brain extraction approach based on FreeSurfer (see Figure 1).

**Figure 1:**
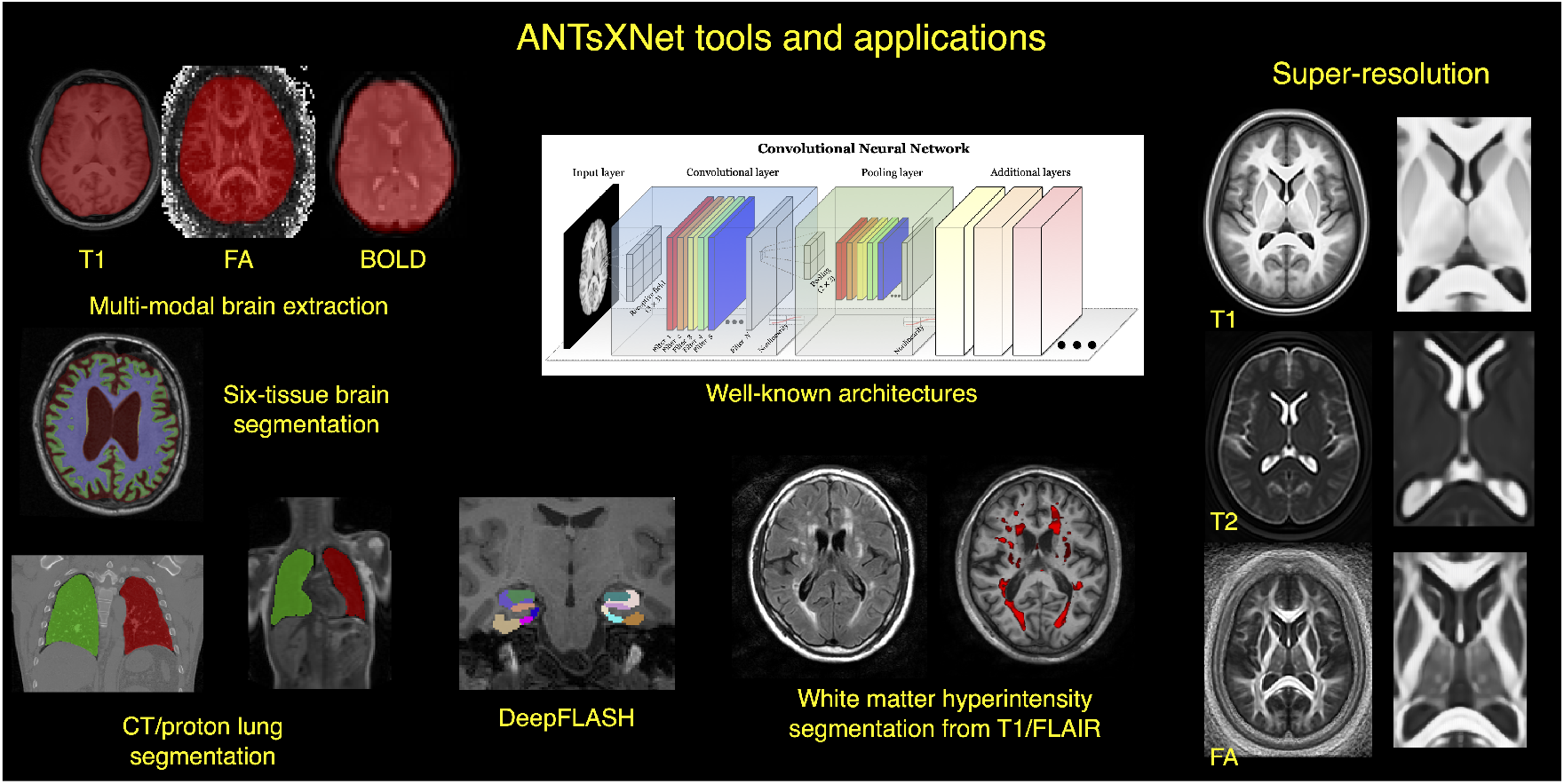
An illustration of the tools and applications available as part of the ANTsRNet and ANTsPyNet deep learning toolkits. Both libraries take advantage of ANTs functionality through their respective language interfaces—ANTsR (R) and ANTsPy (Python). Building on the Keras/TensorFlow language, both libraries standardize popular network architectures within the ANTs ecosystem and are cross-compatible. These networks are used to train models and weights for such applications as brain extraction which are then disseminated to the public.

### The ANTsXNet cortical thickness pipeline

The most recent ANTsX innovation involves the development of deep learning analogs of our popular ANTs cortical thickness cross-sectional^66^ and longitudinal^51^ pipelines within the ANTsXNet framework. Figure 2, adapted from our previous work,^66^ illustrates some of the major changes associated with the single-subject, cross-sectional pipeline. The resulting improvement in efficiency derives primarily from eliminating deformable image registration from the pipeline—a step which has historically been used to propagate prior, population-based information (e.g., tissue maps) to individual subjects for such tasks as brain extraction^58^ and tissue segmentation^68^ which is now configured within the neural networks and trained weights.

**Figure 2:**
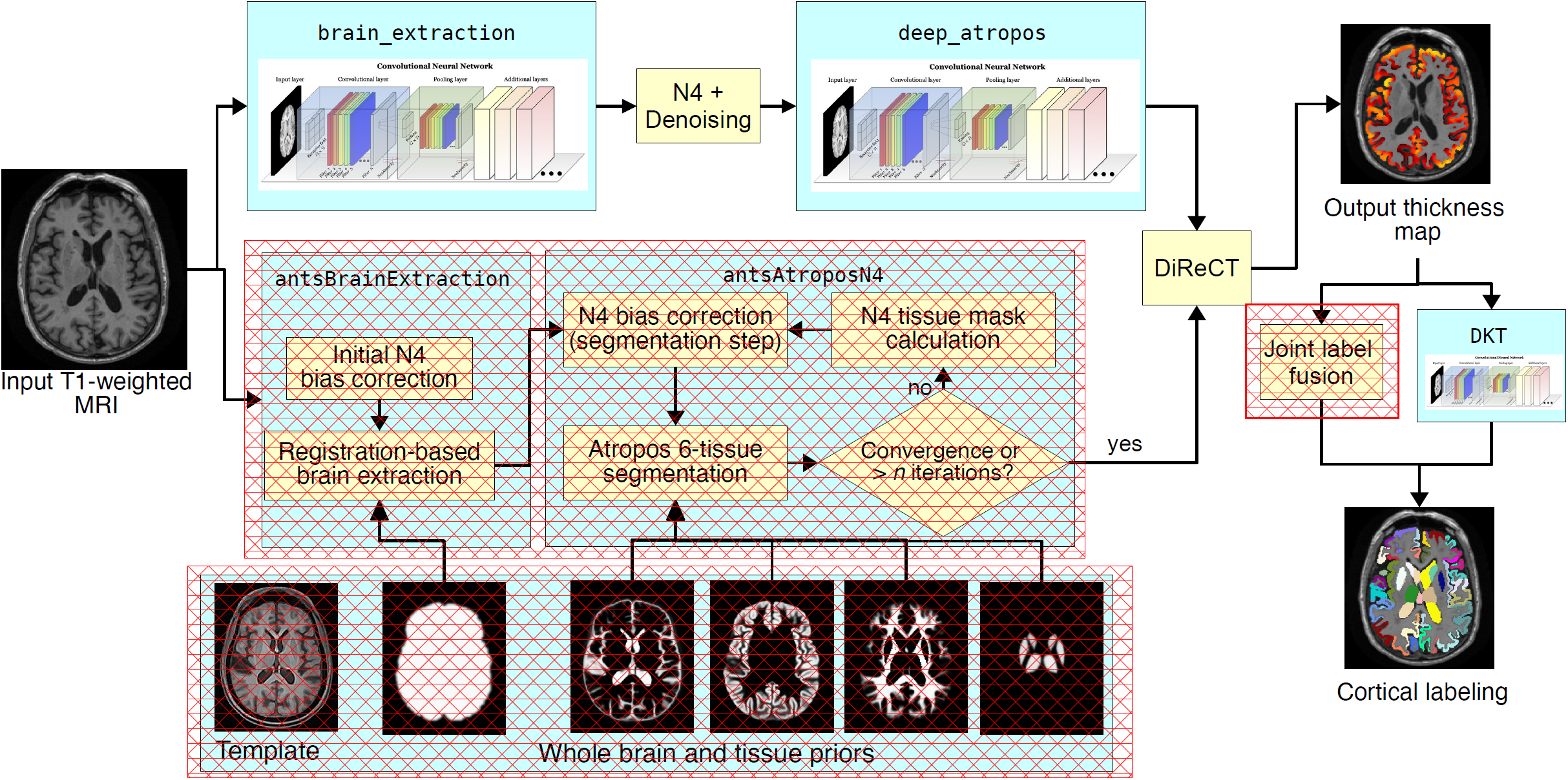
Illustration of the ANTsXNet cortical thickness pipeline and the relationship to its traditional ANTs analog. The hash-designated sections denote pipeline steps which have been obviated by the deep learning approach. These include template-based brain extraction, template-based *n*-tissue segmentation, and joint label fusion for cortical labeling. In our prior work, execution time of the thickness pipeline was dominated by registration. In the deep version of the pipeline, it is dominated by DiReCT. However, we note that registration and DiReCT execute much more quickly than in the past in part due to major improvements in the underlying ITK multi-threading strategy.

These structural MRI processing pipelines are currently available as open-source within the ANTsXNet libraries. Evaluations using both cross-sectional and longitudinal data are described in subsequent sections and couched within the context of our previous publications.^51,66^ Related work has been recently reported by external groups^38,39^ and provides a context for comparison to motivate the utility of the ANTsX ecosystem.

## Results

### Cross-sectional performance evaluation

Due to the absence of ground-truth, we utilize the evaluation strategy from our previous work^66^ where we used cross-validation to build and compare age prediction models from data derived from both the proposed ANTsXNet pipeline and the established ANTs pipeline. Specifically, we use “age” as a well-known and widely-available demographic correlate of cortical thickness^30^ and quantify the predictive capabilities of corresponding random forest classifiers^19^ of the form:

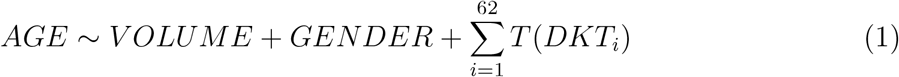

with covariates *GENDER* and *V OLUME* (i.e., total intracranial volume). *T* (*DKT*_*i*_) is the average thickness value in the *i*^*th*^ Desikian-Killiany-Tourville (DKT) region^35^ (cf Table 2). Root mean square error (RMSE) between the actual and predicted ages are the quantity used for comparative evaluation. As we have explained previously,^66^ we find these evaluation measures to be much more useful than other commonly applied criteria as they are closer to assessing the actual utility of these thickness measurements as biomarkers for disease^21^ or growth. In recent work^39^ the authors employ correlation with FreeSurfer thickness values as the primary evaluation for assessing relative performance with ANTs cortical thickness.^66^ This evaluation, unfortunately, is fundamentally flawed in that it is a prime example of a type of circularity analysis^29^ whereby data selection is driven by the same criteria used to evaluate performance. Specifically, the underlying DeepSCAN network used for the tissue segmentation step employs training based on FreeSurfer results which directly influences thickness values as thickness/segmentation are highly correlated and vary characteristically between software packages. Relative performance with ANTs thickness (which does not use FreeSurfer for training) is then assessed by determining correlations with FreeSurfer thickness values. Almost as problematic is their use of repeatability, which they confusingly label as “robustness,” as an additional ranking criterion. Repeatability evaluations should be contextualized within considerations such as the bias-variance tradeoff and quantified using relevant metrics, such as the intra-class correlation coefficient which takes into account both inter- and intra-observer variability.

**Table 2:** The 31 cortical labels (per hemisphere) of the Desikan-Killiany-Tourville atlas. The ROI abbreviations from the R brainGraph package are given in parentheses and used in later figures.

|  |  |
| --- | --- |
| 1) caudal anterior cingulate (**cACC**) | 17) pars orbitalis (**pORB**) |
| 2) caudal middle frontal (**cMFG**) | 18) pars triangularis (**pTRI**) |
| 3) cuneus (**CUN**) | 19) pericalcarine (**periCAL**) |
| 4) entorhinal (**ENT**) | 20) postcentral (**postC**) |
| 5) fusiform (**FUS**) | 21) posterior cingulate (**PCC**) |
| 6) inferior parietal (**IPL**) | 22) precentral (**preC**) |
| 7) inferior temporal (**ITG**) | 23) precuneus (**PCUN**) |
| 8) isthmus cingulate (**iCC**) | 24) rosterior anterior cingulate (**rACC**) |
| 9) lateral occipital (**LOG**) | 25) rostral middle frontal (**rMFG**) |
| 10) lateral orbitofrontal (**LOF**) | 26) superior frontal (**SFG**) |
| 11) lingual (**LING**) | 27) superior parietal (**SPL**) |
| 12) medial orbitofrontal (**MOF**) | 28) superior temporal (**STG**) |
| 13) middle temporal (**MTG**) | 29) supramarginal (**SMAR**) |
| 14) parahippocampal (**PARH**) | 30) transverse temporal (**TT**) |
| 15) paracentral (**paraC**) | 31) insula (**INS**) |
| 16) pars opercularis (**pOPER**) |  |

In addition to the training data listed above, to ensure generalizability, we also compared performance using the SRPB data set^15^ comprising over 1600 participants from 12 sites. Note that we recognize that we are processing a portion of the evaluation data through certain components of the proposed deep learning-based pipeline that were used to train the same pipeline components. Although this does not provide evidence for generalizability (which is why we include the much larger SRPB data set), it is still interesting to examine the results since, in this case, the deep learning training can be considered a type of noise reduction on the final results. It should be noted that training did not use age prediction (or any other evaluation or related measure) as a criterion to be optimized during network model training (i.e., circular analysis).^29^

The results are shown in Figure 3 where we used cross-validation with 500 permutations per model per data set (including a “combined” set) and an 80/20 training/testing split. The ANTsXNet deep learning pipeline outperformed the classical pipeline^66^ in terms of age prediction in all data sets except for IXI. This also includes the cross-validation iteration where all data sets were combined. Additionally, repeatability assessment on the regional cortical thickness values of the MMRR data set yielded ICC values (“average random rater”) of 0.99 for both pipelines.

**Figure 3:**
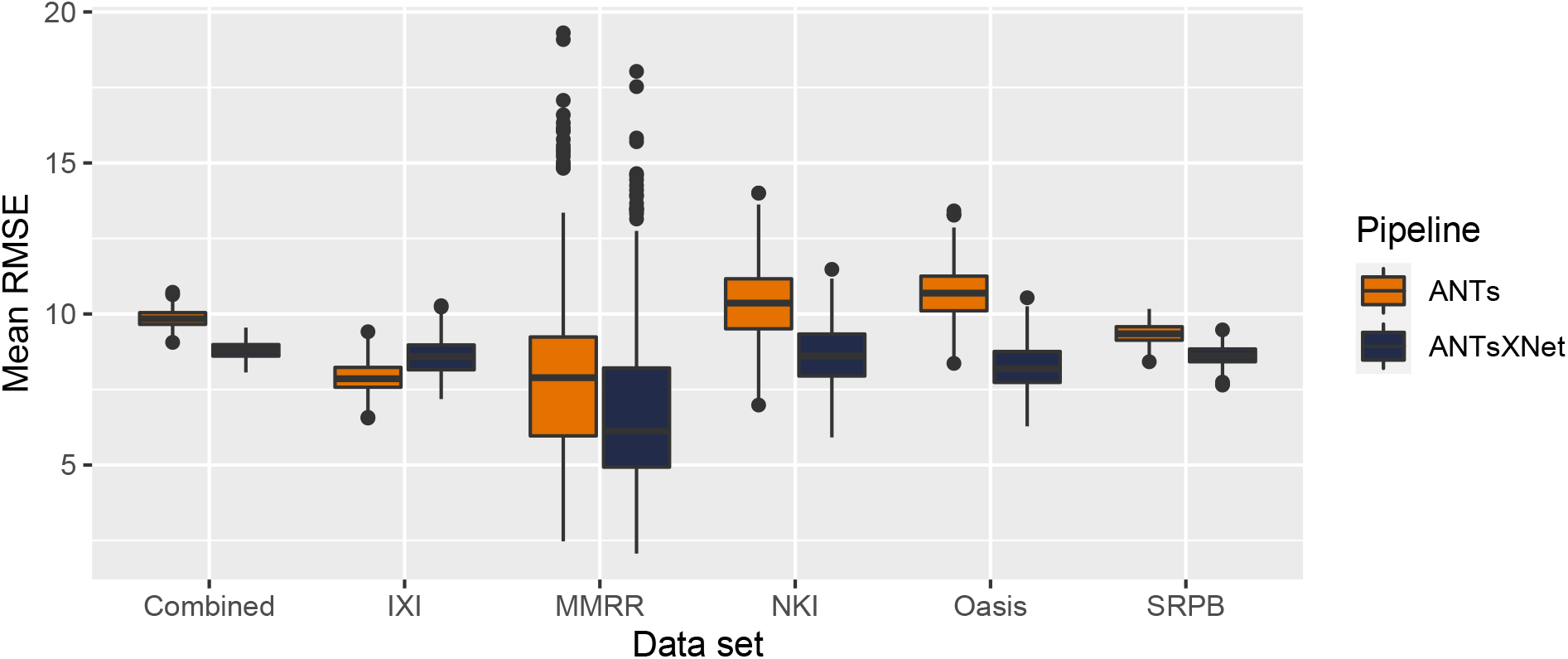
Distribution of mean RMSE values (500 permutations) for age prediction across the different data sets between the traditional ANTs and deep learning-based ANTsXNet pipelines. Total mean values are as follows: Combined—9.3 years (ANTs) and 8.2 years (ANTsXNet); IXI—7.9 years (ANTs) and 8.6 years (ANTsXNet); MMRR—7.9 years (ANTs) and 7.6 years (ANTsXNet); NKI—8.7 years (ANTs) and 7.9 years (ANTsXNet); OASIS—9.2 years (ANTs) and 8.0 years (ANTsXNet); and SRPB—9.2 years (ANTs) and 8.1 years (ANTsXNet).

A comparative illustration of regional thickness measurements between the ANTs and ANTsXNet pipelines is provided in Figure 4 for three different ages spanning the lifespan. Linear models of the form

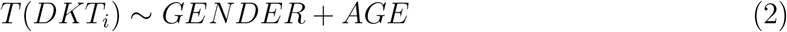

were created for each of the 62 DKT regions for each pipeline. These models were then used to predict thickness values for each gender at ages of 25 years, 50 years, and 75 years and subsequently plotted relative to the absolute maximum predicted thickness value (ANTs: right entorhinal cortex at 25 years, male). Although there appear to be systematic differences between specific regional predicted thickness values (e.g., *T* (*ENT*)_*ANTs*_ *> T* (*ENT*)_*ANTsXNet*_, *T* (*pORB*)_*ANTs*_ *< T* (*pORB*)_*ANTsXNet*_)), a pairwise t-test evidenced no statistically significant difference between the predicted thickness values of the two pipelines.

**Figure 4:**
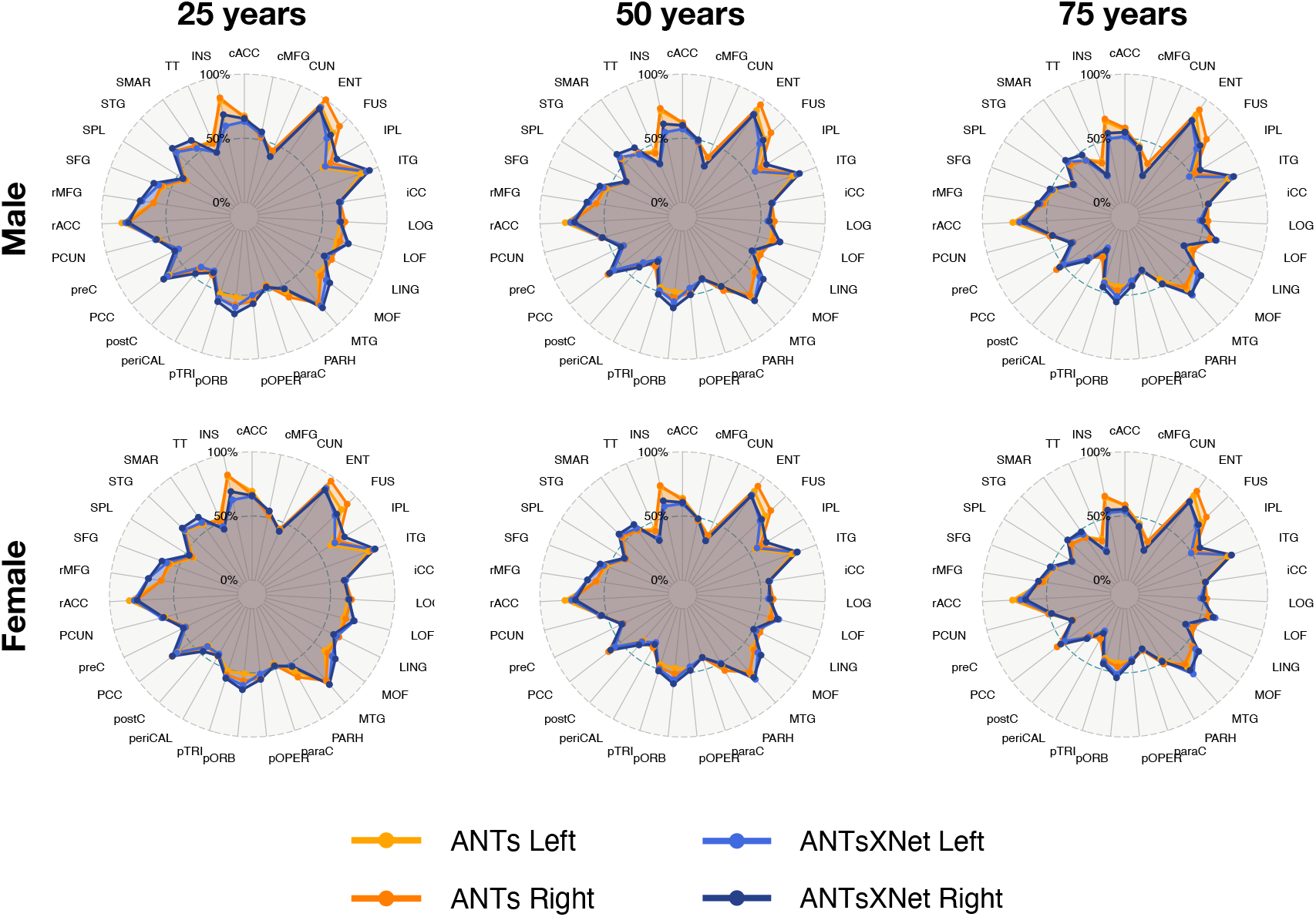
Radar plots enabling comparison of relative thickness values between the ANTs and ANTsXNet cortical thickness pipelines at three different ages sampling the life span. See Table 2 for region abbreviations.

### Longitudinal performance evaluation

Given the excellent performance and superior computational efficiency of the proposed ANTsXNet pipeline for cross-sectional data, we evaluated its performance on longitudinal data using the longitudinally-specific evaluation strategy and data we employed with the introduction of the longitudinal version of the ANTs cortical thickness pipeline.^51^ We also evaluated an ANTsXNet-based pipeline tailored specifically for longitudinal data. In this variant, an SST is generated and processed using the previously described ANTsXNet cross-sectional pipeline which yields tissue spatial priors. These spatial priors are used in our traditional brain segmentation approach^68^. The computational efficiency of this variant is also significantly improved, in part, due to the elimination of the costly SST prior generation which uses multiple registrations combined with joint label fusion.^53^

The ADNI-1 data used for our longitudinal performance evaluation^51^ consists of over 600 subjects (197 cognitive normals, 324 LMCI subjects, and 142 AD subjects) with one or more follow-up image acquisition sessions every 6 months (up to 36 months) for a total of over 2500 images. In addition to the ANTsXNet pipelines (“ANTsXNetCross” and “ANTsXNetLong”) for the current evaluation, our previous work included the FreeSurfer^61^ cross-sectional (“FSCross”) and longitudinal (“FSLong”) streams, the ANTs cross-sectional pipeline (“ANTsCross”) in addition to two longitudinal ANTs-based variants (“ANTsNative” and “ANTsSST”). Two evaluation measurements, one unsupervised and one supervised, were used to assess comparative performance between all seven pipelines. We add the results of the ANTsXNet pipeline cross-sectional and longitudinal evaluations in relation to these other pipelines to provide a comprehensive overview of relative performance.

First, linear mixed-effects (LME)^20^ modeling was used to quantify between-subject and residual variabilities, the ratio of which provides an estimate of the effectiveness of a given biomarker for distinguishing between subpopulations. In order to assess this criteria while accounting for changes that may occur through the passage of time, we used the following Bayesian LME model:

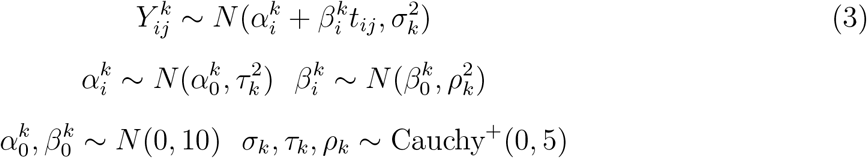

where 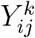 denotes the *i*^*th*^ individual’s cortical thickness measurement corresponding to the *k*^*th*^ region of interest at the time point indexed by *j* and specification of variance priors to half-Cauchy distributions reflects commonly accepted best practice in the context of hierarchical models.^28^ The ratio of interest, *r*^*k*^, per region of the between-subject variability, *τ*_*k*_, and residual variability, *σ*_*k*_ is

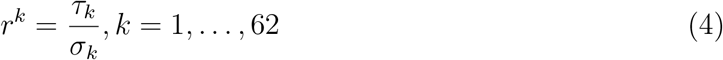

where the posterior distribution of *r*_*k*_ was summarized via the posterior median.

Second, the supervised evaluation employed Tukey post-hoc analyses with false discovery rate (FDR) adjustment to test the significance of the LMCI-CN, AD-LMCI, and AD-CN diagnostic contrasts. This is provided by the following LME model

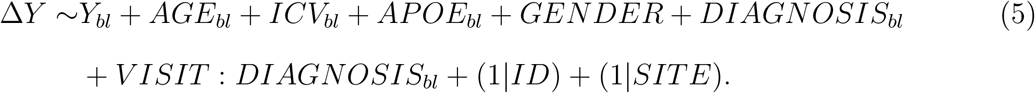

Here, Δ*Y* is the change in thickness of the *k*^*th*^ DKT region from baseline (bl) thickness *Y*_*bl*_ with random intercepts for both the individual subject (*ID*) and the acquisition site. The subject-specific covariates *AGE, APOE* status, *GENDER, DIAGNOSIS, ICV*, and *V ISIT* were taken directly from the ADNIMERGE package.

Results for all pipelines with respect to the longitudinal evaluation criteria are shown in Figures 5 and 6. Figure 5(a) provides the 95% confidence intervals of the variance ratio for all 62 regions of the DKT cortical labeling where ANTsSST consistently performs best with ANTsXNetLong also performing well. These quantities are summarized in Figure 5(b). The second evaluation criteria compares diagnostic differentiation via LMEs. Log p-values are provided in Figure 6 which demonstrate excellent LMCI-CN and AD-CN differentiation for both deep learning pipelines.

**Figure 5:**
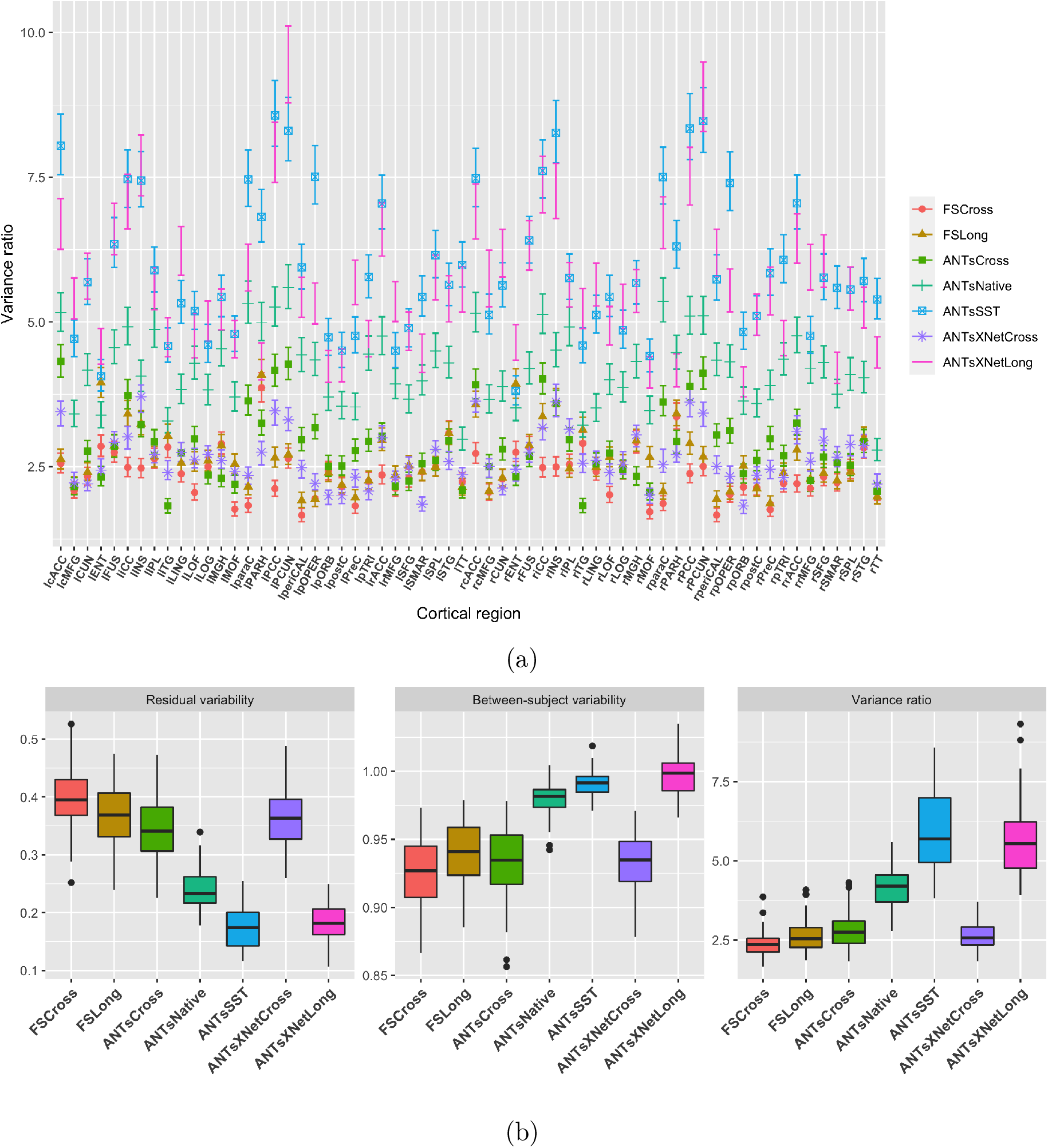
Performance over longitudinal data as determined by the variance ratio. (a) Region-specific 95% confidence intervals of the variance ratio showing the superior performance of the longitudinally tailored ANTsX-based pipelines, including ANTsSST and ANTsXNetLong. (b) Residual variability, between subject, and variance ratio values per pipeline over all DKT regions.

**Figure 6:**
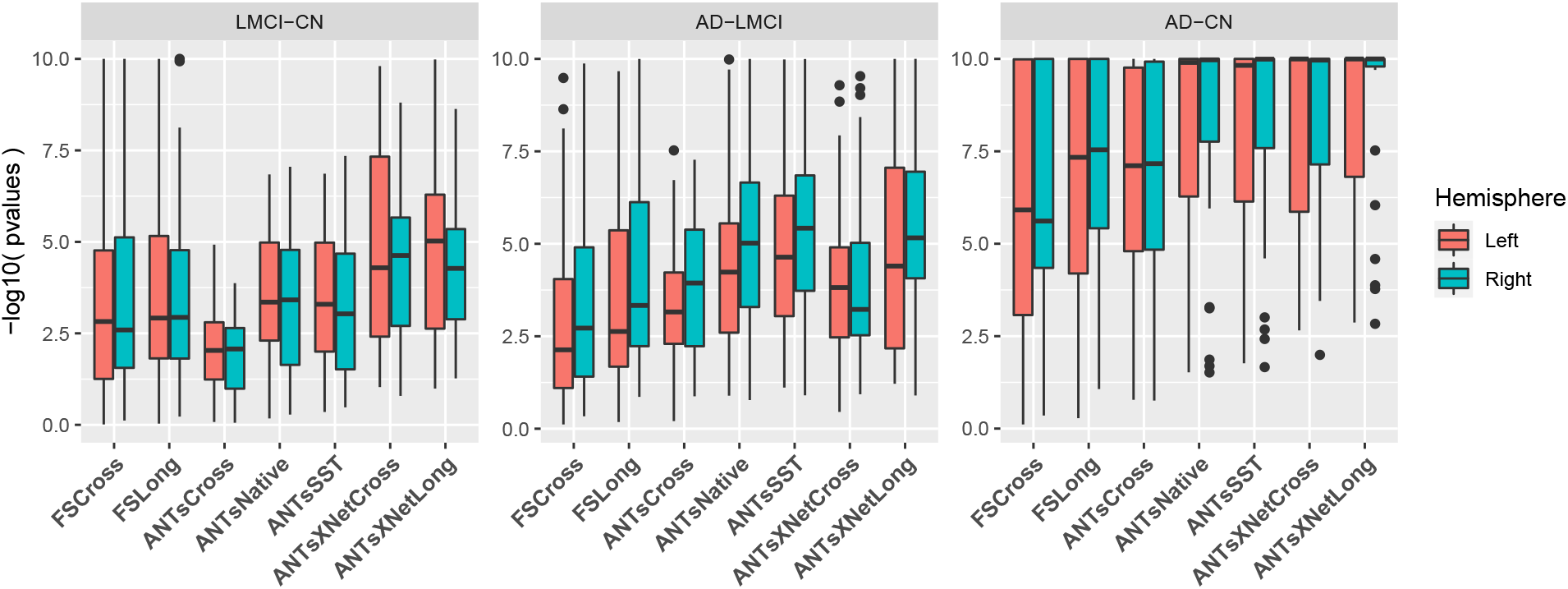
Measures for the supervised evaluation strategy where log p-values for diagnostic differentiation of LMCI-CN, AD-LMCI, and AD-CN subjects are plotted for all pipelines over all DKT regions.

## Discussion

The ANTsX software ecosystem provides a comprehensive framework for quantitative biological and medical imaging. Although ANTs, the original core of ANTsX, is still at the forefront of image registration technology, it has moved significantly beyond its image registration origins. This expansion is not confined to technical contributions (of which there are many) but also consists of facilitating access to a wide range of users who can use ANTsX tools (whether through bash, Python, or R scripting) to construct tailored pipelines for their own studies or to take advantage of our pre-fabricated pipelines. And given the open-source nature of the ANTsX software, usage is not limited, for example, to non-commercial use—a common constraint characteristic of other packages such as the FMRIB Software Library (https://fsl.fmrib.ox.ac.uk/fsl/fslwiki/Licence).

One of our most widely used pipelines is the estimation of cortical thickness from neuroimaging. This is understandable given the widespread usage of regional cortical thickness as a biomarker for developmental or pathological trajectories of the brain. In this work, we used this well-vetted ANTs tool to provide training data for producing alternative variants which leverage deep learning for improved computational efficiency and also provides superior performance with respect to previously proposed evaluation measures for both cross-sectional^66^ and longitudinal scenarios.^51^ In addition to providing the tools which generated the original training data for the proposed ANTsXNet pipeline, the ANTsX ecosystem provides a full-featured platform for the additional steps such as preprocessing (ANTsR/ANTsPy); data augmentation (ANTsR/ANTsPy); network construction and training (ANTsRNet/ANTsPyNet); and visualization and statistical analysis of the results (ANTsR/ANTsPy).

It is the comprehensiveness of ANTsX that provides several advantages over much of the deep learning work that is currently taking place in medical imaging. In other words, various steps in the deep learning training processing (e.g., data augmentation, preprocessing) can all be performed within the same ecosystem where such important details as header information for image geometry are treated the same. In contrast, related work^39^ described and evaluated a similar thickness measurement pipeline. However, due to the lack of a complete processing and analysis framework, training data was generated using the FreeSurfer stream, deep learning-based brain segmentation employed DeepSCAN^27^ (in-house software), and cortical thickness estimation^57^ was generated using the ANTs toolkit. The interested researcher must ensure the consistency of the input/output interface between packages (a task for which the Nipype development team is quite familiar.)

Although potentially advantageous in terms of such issues as computational efficiency and other performance measures, there are a number of limitations associated with the ANTsXNet pipeline that should be mentioned both to guide potential users and possibly motivate future related research. As is the case with many deep learning models, usage is restricted based on training data. For example, much of the publicly available brain data has been anonymized through various defacing protocols. That is certainly the case with the training data used for the ANTsXNet pipeline which has consequences specific to the brain extraction step which could lead to poor performance. We are currently aware of this issue and have provided a temporary workaround while simultaneously resuming training on whole head data to mitigate this issue. Related, although the ANTsXNet pipeline performs relatively well as assessed across lifespan data, performance might be hampered for specific age ranges (e.g., neonates), whereas the traditional ANTs cortical thickness pipeline is more flexible and might provide better age-targeted performance. This is the subject of ongoing research. Additionally, application of the ANTsXNet pipeline would be limited with high-resolution acquisitions. Due to the heavy memory requirements associated with deep learning training, the utility of any resolution greater than 1 mm isotropic would not be leveraged by the existing pipeline. However, there is a potential pipeline variation (akin to the longitudinal variant) that would be worth exploring where Deep Atropos is used only to provide the priors for a subsequent traditional Atropos segmentation on high-resolution data.

In terms of additional future work, the recent surge and utility of deep learning in medical image analysis has significantly guided the areas of active ANTsX development. As demonstrated in this work with our widely used cortical thickness pipelines, there are many potential benefits of deep learning analogs to existing ANTs tools as well as the development of new ones. Performance is mostly comparable-to-superior relative to existing pipelines depending on the evaluation metric. Specifically, the ANTsXNet cross-sectional pipeline does well for the age prediction performance framework and in terms of the ICC. Additionally, this pipeline performs relatively well for longitudinal ADNI data for disease differentiation but not so much in terms of the generic variance ratio criterion. However, for such longitudinal-specific studies, the ANTsXNet longitudinal variant performs well for both performance measures. We see possible additional longitudinal extensions incorporating subject ID and months as additional network inputs.

## Methods

### The original ANTs cortical thickness pipeline

The original ANTs cortical thickness pipeline^66^ consists of the following steps:

- preprocessing: denoising^56^ and bias correction;^67^
- brain extraction;^58^
- brain segmentation with spatial tissue priors^68^ comprising the
  –cerebrospinal fluid (CSF),
  –gray matter (GM),
  –white matter (WM),
  –deep gray matter,
  –cerebellum, and
  –brain stem; and
- cortical thickness estimation.^57^

Our recent longitudinal variant^51^ incorporates an additional step involving the construction of a single subject template (SST)^64^ coupled with the generation of tissue spatial priors of the SST for use with the processing of the individual time points as described above.

Although the resulting thickness maps are conducive to voxel-based^36^ and related analyses^37^, here we employ the well-known Desikan-Killiany-Tourville (DKT)^35^ labeling protocol (31 labels per hemisphere) to parcellate the cortex for averaging thickness values regionally (cf Table 2). This allows us to 1) be consistent in our evaluation strategy for comparison with our previous work^51,66^ and 2) leverage an additional deep learning-based substitution within the proposed pipeline.

### Overview of cortical thickness via ANTsXNet

The entire analysis/evaluation framework, from preprocessing to statistical analysis, is made possible through the ANTsX ecosystem and simplified through the open-source R and Python platforms. Preprocessing, image registration, and cortical thickness estimation are all available through the ANTsPy and ANTsR libraries whereas the deep learning steps are performed through networks constructed and trained via ANTsRNet/ANTsPyNet with data augmentation strategies and other utilities built from ANTsR/ANTsPy functionality.

The brain extraction, brain segmentation, and DKT parcellation deep learning components were trained using data derived from our previous work.^66^ Specifically, the IXI,^18^ MMRR,^31^ NKI,^17^ and OASIS^16^ data sets, and the corresponding derived data, comprising over 1200 subjects from age 4 to 94, were used for network training. Brain extraction employs a traditional 3-D U-net network^44^ with whole brain, template-based data augmentation^32^ whereas brain segmentation and DKT parcellation are processed via 3-D U-net networks with attention gating^33^ on image octant-based batches. Additional network architecture details are given below. We emphasize that a single model (as opposed to ensemble approaches where multiple models are used to produce the final solution)^40^ was created for each of these steps and was used for all the experiments described below.

### Implementation

Software, average DKT regional thickness values for all data sets, and the scripts to perform both the analysis and obtain thickness values for a single subject (cross-sectionally or longitudinally) are provided as open-source. Specifically, all the ANTsX libraries are hosted on GitHub (https://github.com/ANTsX). The cross-sectional data and analysis code are available as .csv files and R scripts at the GitHub repository dedicated to this paper (https://github.com/ntustison/PaperANTsX) whereas the longitudinal data and evaluation scripts are organized with the repository associated with our previous work^51^ (https://githubcom/ntustison/CrossLong).

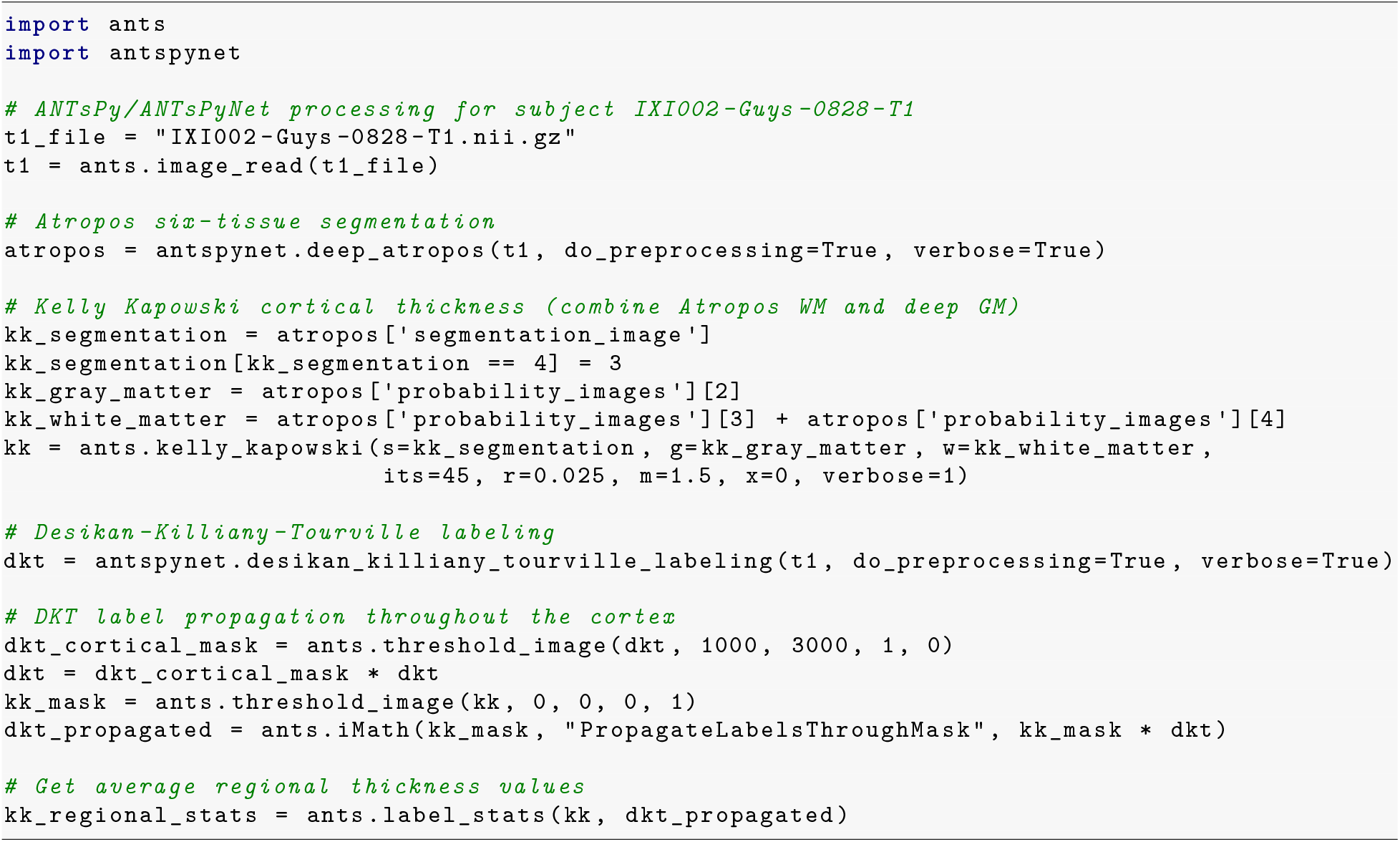

Listing 1: ANTsPy/ANTsPyNet command calls for a single IXI subject in the evaluation study for the cross-sectional pipeline.

In Listing 1 we show the ANTsPy/ANTsPyNet code snippet for cross-sectional processing a single subject which starts with reading the T1-weighted MRI input image, through the generation of the Atropos-style six-tissue segmentation and probability images, application of `ants.kelly_kapowski` (i.e., DiReCT), DKT cortical parcellation, subsequent label propagation through the cortex, and, finally, regional cortical thickness tabulation. The cross-sectional and longitudinal pipelines are encapsulated in the ANTsPyNet functions `antspynet.cortical_thickness` and `antspynet.longitudinal_cortical_thickness,` and re-spectively. Note that there are precise, line-by-line R-based analogs available through ANTsR/ANTsRNet.

Both the `ants.deep_atropos` and of `antspynet.desikan_killiany_tourville_labeling` functions perform brain extraction using the `antspynet.brain_extraction` function. Internally, `antspynet.brain_extraction` contains the requisite code to build the network and assign the appropriate hyperparameters. The model weights are automatically downloaded from the online hosting site https://figshare.com (see the function `get_pretrained_network` in ANTsPyNet or getPretrainedNetwork in ANTsRNet for links to all models and weights) and loaded to the constructed network. `antspynet.brain_extraction` performs a quick translation transformation to a specific template (also downloaded automatically) using the centers of intensity mass, a common alignment initialization strategy. This is to ensure proper gross orientation. Following brain extraction, preprocessing for the other two deep learning components includes `ants.denoise_image` and `ants.n4_bias_correction` and an affine-based reorientation to a version of the MNI template.^34^

We recognize the presence of some redundancy due to the repeated application of certain preprocessing steps. Thus, each function has a `do_preprocessing` option to eliminate this redundancy for knowledgeable users but, for simplicity in presentation purposes, we do not provide this modified pipeline here. Although it should be noted that the time difference is minimal considering the longer time required by ants.kelly_kapowski. ants.deep_atropos returns the segmentation image as well as the posterior probability maps for each tissue type listed previously. `antspynet.desikan_killiany_tourville_labeling` returns only the segmentation label image which includes not only the 62 cortical labels but the remaining labels as well. The label numbers and corresponding structure names are given in the program description/help. Because the DKT parcellation will, in general, not exactly coincide with the non-zero voxels of the resulting cortical thickness maps, we perform a label propagation step to ensure the entire cortex, and only the non-zero thickness values in the cortex, are included in the tabulated regional values.

As mentioned previously, the longitudinal version, `antspynet.longitudinal_cortical_thickness,` adds an SST generation step which can either be provided as a program input or it can be constructed from spatial normalization of all time points to a specified template. `ants.deep_atropos` is applied to the SST yielding spatial tissues priors which are then used as input to `ants.atropos` for each time point. `ants.kelly_kapowski` is applied to the result to generate the desired cortical thickness maps.

Computational time on a CPU-only platform is approximately 1 hour primarily due to `ants.kelly_kapowski` processing. Other preprocessing steps, i.e., bias correction and denoising, are on the order of a couple minutes. This total time should be compared with 4 − 5 hours using the traditional pipeline employing the `quick` registration option or 10 − 15 hours with the more comprehensive registration parameters employed). As mentioned previously, elimination of the registration-based propagation of prior probability images to individual subjects is the principal source of reduced computational time. For ROI-based analyses, this is in addition to the elimination of the optional generation of a population-specific template. Additionally, the use of `antspynet.desikan_killiany_tourville_labeling,` for cortical labeling (which completes in less than five minutes) eliminates the need for joint label fusion which requires multiple pairwise registrations for each subject in addition to the fusion algorithm itself.

### Training details

Training differed slightly between models and so we provide details for each of these components below. For all training, we used ANTsRNet scripts and custom batch generators. Although the network construction and other functionality is available in both ANTsPyNet and ANTsRNet (as is model weights compatibility), we have not written such custom batch generators for the former (although this is on our to-do list). In terms of hardware, all training was done on a DGX (GPUs: 4X Tesla V100, system memory: 256 GB LRDIMM DDR4).

### T1-weighted brain extraction

A whole-image 3-D U-net model^44^ was used in conjunction with multiple training sessions employing a Dice loss function followed by categorical cross entropy. Training data was derived from the same multi-site data described previously processed through our registration-based approach.^58^ A center-of-mass-based transformation to a standard template was used to standardize such parameters as orientation and voxel size. However, to account for possible different header orientations of input data, a template-based data augmentation scheme was used^32^ whereby forward and inverse transforms are used to randomly warp batch images between members of the training population (followed by reorientation to the standard template). A digital random coin flipping for possible histogram matching^26^ between source and target images further increased data augmentation. The output of the network is a probabilistic mask of the brain. The architecture consists of four encoding/decoding layers with eight filters at the base layer which doubled every layer. Although not detailed here, training for brain extraction in other modalities was performed similarly.

### Deep Atropos

Dealing with 3-D data presents unique barriers for training that are often unique to medical imaging. Various strategies are employed such as minimizing the number of layers and/or the number of filters at the base layer of the U-net architecture (as we do for brian extraction). However, we found this to be too limiting for capturing certain brain structures such as the cortex. 2-D and 2.5-D approaches are often used with varying levels of success but we also found better performance using full 3-D information. This led us to try randomly selected 3-D patches of various sizes. However, for both the six-tissue segmentations and DKT parcellations, we found that an octant-based patch strategy yielded the desired results. Specifically, after a brain extracted affine normalization to the MNI template, the normalized image is cropped to a size of [160, 190, 160]. Overlapping octant patches of size [112, 112, 112] were extracted from each image and trained using a batch size of 12 such octant patches with weighted categorical cross entropy as the loss function. The architecture consists of four encoding/decoding layers with 16 filters at the base layer which doubled every layer.

As we point out in our earlier work,^66^ obtaining proper brain segmentation is perhaps the most critical step to estimating thickness values that have the greatest utility as a potential biomarker. In fact, the first and last authors (NT and BA, respectively) spent much time during the original ANTs pipeline development^66^ trying to get the segmentation correct which required manually looking at many images and manually adjusting where necessary. This fine-tuning is often omitted or not considered when other groups^24,25,39^ use components of our cortical thickness pipeline which can be potentially problematic^70^. Fine-tuning for this particular workflow was also performed between the first and last authors using manual variation of the weights in the weighted categorical cross entropy. Specifically, the weights of each tissue type were altered in order to produce segmentations which most resemble the traditional Atropos segmentations. Ultimately, we settled on a weight vector of (0.05, 1.5, 1, 3, 4, 3, 3) for the CSF, GM, WM, Deep GM, brain stem, and cerebellum, respectively. Other hyperparameters can be directly inferred from explicit specification in the actual code. As mentioned previously, training data was derived from application of the ANTs Atropos segmentation^68^ during the course of our previous work.^66^ Data augmentation included small affine and deformable perturbations using `antspynet.randomly_transform_image_data` and random contralateral flips.

### Desikan-Killiany-Tourville parcellation

Preprocessing for the DKT parcellation training was similar to the Deep Atropos training. However, the number of labels and the complexity of the parcellation required deviation from other training steps. First, labeling was split into an inner set and an outer set. Subsequent training was performed separately for both of these sets. For the cortical labels, a set of corresponding input prior probability maps were constructed from the training data (and are also available and automatically downloaded, when needed, from https://figshare.com). Training occurred over multiple sessions where, initially, categorical cross entropy was used and then subsquently refined using a Dice loss function. Whole-brain training was performed on a brain-cropped template size of [96, 112, 96]. Inner label training was performed similarly to our brain extraction training where the number of layers at the base layer was reduced to eight. Training also occurred over multiple sessions where, initially, categorical cross entropy was used and then subsquently refined using a Dice loss function. Other hyperparameters can be directly inferred from explicit specification in the actual code. Training data was derived from application of joint label fusion^63^ during the course of our previous work.^66^ When calling `antspynet.desikan_killiany_tourville_labeling,` inner labels are estimated first followed by the outer cortical labels.

### Other softwares

Several R^1^ packages were used in preparation of this manuscript including R Markdown,^10–12^ lme4,^7^ RStan,^6^ ggplot2,^9^ and ggradar2.^8^ Other packages used include Apple Pages,^3^ ITK-SNAP,^2^ LibreOffice,^4^ and diagrams.net.^5^

## Data Availability

All imaging data is available from public repositories. All software, scripts, and tabulated measurements used to produce the figures and other results are in the specified GitHub repositories.

https://github.com/ntustison/PaperANTsX/

https://github.com/ntustison/CrossLong

## Acknowledgments

Support for the research reported in this work includes funding from the National Heart, Lung, and Blood Institute of the National Institutes of Health (R01HL133889) and a combined grant from Cohen Veterans Bioscience (CVB-461) and the Office of Naval Research (N00014-18-1-2440).

Data used in preparation of this article were obtained from the Alzheimer’s Disease Neuroimaging Initiative (ADNI) database (http://adni.loni.usc.edu). As such, the investigators within the ADNI contributed to the design and implementation of ADNI and/or provided data but did not participate in analysis or writing of this report. A complete listing of ADNI investigators can be found at: http://adni.loni.usc.edu/wp-content/uploads/how to apply/AD NI Acknowledgement List.pdf

Data collection and sharing for this project was funded by the Alzheimer’s Disease Neuroimaging Initiative (ADNI) (National Institutes of Health Grant U01 AG024904) and DOD ADNI (Department of Defense award number W81XWH-12-2-0012). ADNI is funded by the National Institute on Aging, the National Institute of Biomedical Imaging and Bioengineering, and through generous contributions from the following: AbbVie, Alzheimer’s Association; Alzheimer’s Drug Discovery Foundation; Araclon Biotech; BioClinica, Inc.; Biogen; Bristol-Myers Squibb Company; CereSpir, Inc.; Cogstate; Eisai Inc.; Elan Pharmaceuticals, Inc.; Eli Lilly and Company; EuroImmun; F. Hoffmann-La Roche Ltd and its affiliated company Genentech, Inc.; Fujirebio; GE Healthcare; IXICO Ltd.; Janssen Alzheimer Immunotherapy Research & Development, LLC.; Johnson & Johnson Pharmaceutical Research & Development LLC.; Lumosity; Lundbeck; Merck & Co., Inc.; Meso Scale Diagnostics, LLC.; NeuroRx Research; Neurotrack Technologies; Novartis Pharmaceuticals Corporation; Pfizer Inc.; Piramal Imaging; Servier; Takeda Pharmaceutical Company; and Transition Therapeutics. The Canadian Institutes of Health Research is providing funds to support ADNI clinical sites in Canada. Private sector contributions are facilitated by the Foundation for the National Institutes of Health (www.fnih.org). The grantee organization is the Northern California Institute for Research and Education, and the study is coordinated by the Alzheimer’s Therapeutic Research Institute at the University of Southern California. ADNI data are disseminated by the Laboratory for Neuro Imaging at the University of Southern California.

## Author contributions

- Conception and design N.T., A.H., M.Y., J.S., B.A.
- Analysis and interpretation N.T., A.H., D.G., M.Y., J.S. B.A.
- Creation of new software N.T., P.C., H.J., J.M., G.D., J.D., S.D., N.C., J.G., B.A.
- Drafting of manuscript N.T., A.H., P.C., H.J., J.M., G.D., J.G., B.A.

## References

1. R Core Team. R: A language and environment for statistical computing. (R Foundation for Statistical Computing, 2020).

2. Yushkevich, P. A. et al.. User-guided 3D active contour segmentation of anatomical structures: Significantly improved efficiency and reliability. Neuroimage 31, 1116–1128 (2006).

3. https://www.apple.com/pages/.

4. https://www.libreoffice.org/.

5. https://app.diagrams.net.

6. Stan Development Team. RStan: The R interface to Stan. (2020).

7. Bates, D., Mächler, M., Bolker, B. & Walker, S. Fitting linear mixed-effects models using lme4. Journal of Statistical Software 67, 1–48 (2015).

8. https://github.com/xl0418/ggradar2.

9. Wickham, H. ggplot2: Elegant graphics for data analysis. (Springer-Verlag New York, 2016).

10. Allaire, J. et al. Rmarkdown: Dynamic documents for r. (2021).

11. Xie, Y., Allaire, J. J. & Grolemund, G. R markdown: The definitive guide. (Chapman; Hall/CRC, 2018).

12. Xie, Y., Dervieux, C. & Riederer, E. R markdown cookbook. (Chapman; Hall/CRC, 2020).

13. Fu, Y. et al. DeepReg: A deep learning toolkit for medical image registration. Journal of Open Source Software 5, 2705 (2020).

14. Vos, B. D. de et al. A deep learning framework for unsupervised affine and deformable image registration. Med Image Anal 52, 128–143 (2019).

15. https://bicr-resource.atr.jp/srpbs1600/.

16. https://www.oasis-brains.org.

17. http://fcon_1000.projects.nitrc.org/indi/pro/nki.html.

18. https://brain-development.org/ixi-dataset/.

19. Breiman, L. Random forests. Machine Learning 45, 5–32 (2001).

20. Verbeke, G. Linear mixed models for longitudinal data. in Linear mixed models in practice 63–153 (Springer, 1997).

21. Holbrook, A. J. et al. Anterolateral entorhinal cortex thickness as a new biomarker for early detection of Alzheimer’s disease. Alzheimer’s & Dementia: Diagnosis, Assessment & Disease Monitoring 12, e12068 (2020).

22. Haris, M., Shakhnarovich, G. & Ukita, N. Deep back-projection networks for super-resolution. in 2018 IEEE/CVF Conference on Computer Vision and Pattern Recognition 1664–1673 (2018). doi:10.1109/CVPR.2018.00179.

23. Bashyam, V. M. et al. MRI signatures of brain age and disease over the lifespan based on a deep brain network and 14,468 individuals worldwide. Brain 143, 2312–2324 (2020).

24. Clarkson, M. J. et al. A comparison of voxel and surface based cortical thickness estimation methods. Neuroimage 57, 856–65 (2011).

25. Schwarz, C. G. et al. A large-scale comparison of cortical thickness and volume methods for measuring alzheimer’s disease severity. Neuroimage Clin 11, 802–812 (2016).

26. Nyúl, L. G. & Udupa, J. K. On standardizing the MR image intensity scale. Magn Reson Med 42, 1072–81 (1999).

27. McKinley, R. et al. Few-shot brain segmentation from weakly labeled data with deep heteroscedastic multi-task networks. CoRR abs/1904.02436, (2019).

28. Gelman, A. & others. Prior distributions for variance parameters in hierarchical models (comment on article by Browne and Draper). Bayesian analysis 1, 515–534 (2006).

29. Kriegeskorte, N., Simmons, W. K., Bellgowan, P. S. F. & Baker, C. I. Circular analysis in systems neuroscience: The dangers of double dipping. Nat Neurosci 12, 535–40 (2009).

30. Lemaitre, H. et al. Normal age-related brain morphometric changes: Nonuniformity across cortical thickness, surface area and gray matter volume? Neurobiol Aging 33, 617.e1–9 (2012).

31. Landman, B. A. et al. Multi-parametric neuroimaging reproducibility: A 3-T resource study. Neuroimage 54, 2854–66 (2011).

32. Tustison, N. J. et al. Convolutional neural networks with template-based data augmentation for functional lung image quantification. Acad Radiol 26, 412–423 (2019).

33. Schlemper, J. et al. Attention gated networks: Learning to leverage salient regions in medical images. Med Image Anal 53, 197–207 (2019).

34. Fonov, V. S., Evans, A. C., McKinstry, R. C., Almli, C. & Collins, D. L. Unbiased nonlinear average age-appropriate brain templates from birth to adulthood. NeuroImage S102, (2009).

35. Klein, A. & Tourville, J. 101 labeled brain images and a consistent human cortical labeling protocol. Front Neurosci 6, 171 (2012).

36. Ashburner, J. & Friston, K. J. Voxel-based morphometry–the methods. Neuroimage 11, 805–21 (2000).

37. Avants, B. et al. Eigenanatomy improves detection power for longitudinal cortical change. Med Image Comput Comput Assist Interv 15, 206–13 (2012).

38. Henschel, L. et al. FastSurfer - a fast and accurate deep learning based neuroimaging pipeline. Neuroimage 219, 117012 (2020).

39. Rebsamen, M., Rummel, C., Reyes, M., Wiest, R. & McKinley, R. Direct cortical thickness estimation using deep learning-based anatomy segmentation and cortex parcellation. Hum Brain Mapp (2020) doi:10.1002/hbm.25159.

40. Li, H. et al. Fully convolutional network ensembles for white matter hyperintensities segmentation in MR images. Neuroimage 183, 650–665 (2018).

41. Tustison, N. J., Avants, B. B. & Gee, J. C. Learning image-based spatial transformations via convolutional neural networks: A review. Magn Reson Imaging 64, 142–153 (2019).

42. Balakrishnan, G., Zhao, A., Sabuncu, M. R., Guttag, J. & Dalca, A. V. VoxelMorph: A learning framework for deformable medical image registration. IEEE Trans Med Imaging (2019) doi:10.1109/TMI.2019.2897538.

43. Goubran, M. et al. Hippocampal segmentation for brains with extensive atrophy using three-dimensional convolutional neural networks. Hum Brain Mapp 41, 291–308 (2020).

44. Falk, T. et al. U-net: Deep learning for cell counting, detection, and morphometry. Nat Methods 16, 67–70 (2019).

45. Gorgolewski, K. et al. Nipype: A flexible, lightweight and extensible neuroimaging data processing framework in python. Front Neuroinform 5, 13 (2011).

46. Muschelli, J. et al. Neuroconductor: An R platform for medical imaging analysis. Biostatistics 20, 218–239 (2019).

47. Halchenko, Y. O. & Hanke, M. Open is not enough. Let’s take the next step: An integrated, community-driven computing platform for neuroscience. Front Neuroinform 6, 22 (2012).

48. Gorgolewski, K. J. et al. The brain imaging data structure, a format for organizing and describing outputs of neuroimaging experiments. Sci Data 3, 160044 (2016).

49. De Leener, B. et al. SCT: Spinal cord toolbox, an open-source software for processing spinal cord MRI data. Neuroimage 145, 24–43 (2017).

50. Esteban, O. et al. fMRIPrep: A robust preprocessing pipeline for functional MRI. Nat Methods 16, 111–116 (2019).

51. Tustison, N. J. et al. Longitudinal mapping of cortical thickness measurements: An Alzheimer’s Disease Neuroimaging Initiative-based evaluation study. J Alzheimers Dis (2019) doi:10.3233/JAD-190283.

52. Tustison, N. J. & Gee, J. C. N4ITK: Nick’s N3 ITK implementation for MRI bias field correction. The Insight Journal (2009).

53. Wang, H. et al. Multi-atlas segmentation with joint label fusion. IEEE Trans Pattern Anal Mach Intell 35, 611–23 (2013).

54. Bajcsy, R. & Kovacic, S. Multiresolution elastic matching. Computer Vision, Graphics, and Image Processing 46, 1–21 (1989).

55. Bajcsy, R. & Broit, C. Matching of deformed images. in Sixth International Conference on Pattern Recognition (ICPR’82) 351–353 (1982).

56. Manjón, J. V., Coupé, P., Martí-Bonmatí, L., Collins, D. L. & Robles, M. Adaptive non-local means denoising of MR images with spatially varying noise levels. J Magn Reson Imaging 31, 192–203 (2010).

57. Das, S. R., Avants, B. B., Grossman, M. & Gee, J. C. Registration based cortical thickness measurement. Neuroimage 45, 867–79 (2009).

58. Avants, B. B., Klein, A., Tustison, N. J., Woo, J. & Gee, J. C. Evaluation of open-access, automated brain extraction methods on multi-site multi-disorder data. in 16th annual meeting for the organization of human brain mapping (2010).

59. Gee, J., Sundaram, T., Hasegawa, I., Uematsu, H. & Hatabu, H. Characterization of regional pulmonary mechanics from serial magnetic resonance imaging data. Acad Radiol 10, 1147–52 (2003).

60. Avants, B. B., Epstein, C. L., Grossman, M. & Gee, J. C. Symmetric diffeomorphic image registration with cross-correlation: Evaluating automated labeling of elderly and neurodegenerative brain. Med Image Anal 12, 26–41 (2008).

61. Fischl, B. FreeSurfer. Neuroimage 62, 774–81 (2012).

62. Menze, B., Reyes, M. & Van Leemput, K. The multimodal brain tumor image segmentation benchmark (BRATS). IEEE Trans Med Imaging (2014) doi:10.1109/TMI.2014.2377694.

63. Wang, H. & Yushkevich, P. A. Multi-atlas segmentation with joint label fusion and corrective learning-an open source implementation. Front Neuroinform 7, 27 (2013).

64. Avants, B. B. et al. The optimal template effect in hippocampus studies of diseased populations. Neuroimage 49, 2457–66 (2010).

65. Klein, A. et al. Evaluation of 14 nonlinear deformation algorithms applied to human brain MRI registration. Neuroimage 46, 786–802 (2009).

66. Tustison, N. J. et al. Large-scale evaluation of ANTs and FreeSurfer cortical thickness measurements. Neuroimage 99, 166–79 (2014).

67. Tustison, N. J. et al. N4ITK: Improved N3 bias correction. IEEE Trans Med Imaging 29, 1310–20 (2010).

68. Avants, B. B., Tustison, N. J., Wu, J., Cook, P. A. & Gee, J. C. An open source multivariate framework for n-tissue segmentation with evaluation on public data. Neuroinformatics 9, 381–400 (2011).

69. Murphy, K. et al. Evaluation of registration methods on thoracic CT: The EMPIRE10 challenge. IEEE Trans Med Imaging 30, 1901–20 (2011).

70. Tustison, N. J. et al. Instrumentation bias in the use and evaluation of scientific software: Recommendations for reproducible practices in the computational sciences. Front Neurosci 7, 162 (2013).

